# Validation of Digital Pathology Platform for Metabolic-Associated Steatohepatitis for Clinical Trials

**DOI:** 10.1101/2023.09.01.23294940

**Authors:** Hanna Pulaski, Shraddha S. Mehta, Laryssa C. Manigat, Stephanie Kaufman, Hypatia Hou, ILKe Nalbantoglu, Xuchen Zhang, Emily Curl, Ross Taliano, Tae Hun Kim, Michael Torbenson, Jonathan N Glickman, Murray B Resnick, Neel Patel, Cristin E. Taylor, Pierre Bedossa, Michael C Montalto, Andrew H Beck, Katy E Wack

## Abstract

**Aims:** Determine if pathologic assessment of disease activity in steatohepatitis, performed using Whole Slide Images (WSIs) on the AISight Clinical Trials platform, yields results that are comparable to those obtained from the analysis performed using glass slides.

**Methods and Results:** The accuracy of scoring for steatohepatitis (NAS ≥4 with ≥1 for each feature and absence of atypical features suggestive of other liver disease) performed on the WSI viewing platform was evaluated against scoring conducted on glass slides. Both methods were assessed for overall percent agreement (OPA) with a consensus ‘ground truth’ (GT) score, defined as the median score of a panel of 3 expert pathologists on glass slides. Each case was also read by 3 different pathologists, once on glass and once using WSIs with a minimum 2-week washout period between glass and WSI reads. It was demonstrated that the average OPA across 3 pathologists of WSI scoring with GT was non-inferior to the average OPA of glass scoring with GT (non-inferiority margin of -0.05, difference of -0.001, 95% CI of (−0.027,0.026), and p<0.0001). For each pathologist, there was a similar average OPA of WSI and glass reads with glass GT (pathologist A 0.843 and 0.849, pathologist B 0.633 and 0.605 and pathologist C 0.755 and 0.780), with intra-reader, inter-modality agreements per histologic feature being greater than published intra-reader agreements.

**Conclusion:** Accuracy of digital reads for steatohepatitis using WSIs is equivalent to glass reads in the context of a clinical trial for scoring using the Clinical Research Network scoring system.

## Introduction

Metabolic dysfunction-associated steatotic liver disease (MASLD; formerly referred to as nonalcoholic fatty liver disease or NAFLD) is rising in prevalence globally, with an estimated 25% of the world’s population affected (1). Due to this increased burden of disease, liver decompensation due to progression of Metabolic-Associated Steatohepatitis (MASH; formerly referred to as nonalcoholic steatohepatitis or NASH) is the leading cause of liver transplant in women (2) and expected to become the overall leading cause of liver transplant (3). There are currently no approved therapies for MASH and there is a large unmet need for clinical intervention in this patient population. A large number of clinical trials are ongoing to identify therapies for MASH, with changes in histologic features as the primary endpoint for most of these trials. Due to the variation between expert pathologists, regulatory bodies are recommending multiple or consensus reads to reduce individual bias and increase quality and consistency. However, obtaining the evaluations from several expert pathologists for the same participant on glass slides for enrollment and follow-up time point reads is challenging, as shipping the slides around the country or in some cases around the world is time consuming and comes with the hazards of slide breakage during the shipment.

Currently, the practice of pathology is increasingly adopting and incorporating digital pathology into clinical workflows. Numerous studies have shown high accuracy for providing primary diagnoses using Whole Slide Images (WSIs) of glass slides (4–14). The current gold standard to establish the diagnosis of MASH is histopathologic analysis of a liver biopsy. The diagnosis is established by the presence of a characteristic histologic pattern of ≥5% hepatic steatosis, lobular inflammation, and hepatocellular ballooning in the appropriate clinical setting and provided other potential cases of metabolic-associated liver disease such as significant alcohol intake have been excluded. In 1999, a semi-quantitative grading and staging system to describe and unify the approach of pathologists to the histopathologic lesions of MASH was proposed by Brunt et al (15). A semi-quantitative activity grade (NAFLD activity score, or NAS) was assigned by a combination of parameters including steatosis, lobular inflammation, and hepatocyte ballooning. NAS ≥4 is used in many clinical trials as a definition of steatohepatitis for enrollment criteria (16–21) and, because the WSI viewing platform is utilized in the context of clinical trials, we chose to define steatohepatitis as NAS ≥4 with a score of ≥1 for each feature and absence of atypical features suggestive of other liver disease.

Importantly, as a secondary analysis, we determined Linearly Weighted Kappa (WK) concordance for intra-reader, inter-modality assessment of steatosis, lobular inflammation, hepatocellular ballooning, fibrosis and overall NAS. Exploratory analysis compared the score-based accuracy of assessing steatosis, hepatocellular ballooning, lobular inflammation and fibrosis for expert pathologist readers evaluating on glass and digital compared to an independent consensus ground truth (GT). Per this context of use, a study population was chosen which represents MASH trial screening and enrolled populations from multiple, completed trials as well as commercially available clinical non-MASH samples. This population was enriched for borderline MASLD/steatohepatitis cases along with non-MASH cases, in order to thoroughly assess the ability to use digital pathology as a surrogate for glass reads in MASH trials.

## Materials and Methods

### Ethics

This study was approved by the WCG IRB (IRB00000533).

### Digital Pathology Platform

The AISight Clinical Trials platform (v3.3.1) is a Good Clinical Practice (GCP) compliant research use only cloud-based software as a service that serves as an interface for viewing WSIs and algorithm outputs. Platform configurability allows for maximum flexibility in leveraging digital pathology to improve subject outcomes in clinical research. All pathologists performing digital reads in this study were trained on the use of the platform and completed practice cases prior to study start. The 3 pathologists using the WSI viewing platform had 4, 6 and 8 years of digital pathology experience.

### Case selection and scanning

Existing de-identified glass slides from a third-party vendor and from completed clinical trials (screen failures and enrolled cases) were utilized in this study. Each case in this study had 2 slides – H&E and Masson’s trichrome. Slides were first scanned at a College of American Pathologists (CAP) accredited, Clinical Laboratory Improvement Amendments (CLIA)-certified lab on Leica Aperio AT2 scanner at 40x magnification, uploaded to WSI viewing platform after image quality control and then distributed for glass reads.

The slide set consisted of 160 cases from liver needle biopsies. Two thirds of the cases were chosen from patients with steatohepatitis (defined as NAS ≥4 with a score of ≥1 for each feature and absence of atypical features suggestive of other liver disease) based on the original trial central pathology scores, and the remaining 1/3 were from MASLD and other liver disorders encountered during clinical trial screening and follow-up timepoints (for inclusion of atypical features suggestive of other liver disease). Based on the original trial individual central pathologist scores, 5-10% of the cases were chosen to be diagnostically challenging or borderline steatohepatitis. This borderline category was defined as NAS ≥4 with a score of 0 for at least one of the histologic features (steatosis, lobular inflammation, and hepatocellular ballooning), NAS =4 with a score of ≥1 for each of the features or NAS =3. For glass reads, the 160 cases were split into 3 batches and the pathologists read 1 batch at a time (all WSIs read first and glass slides read after a minimum of 2-week washout).

### Pathologists’ reads

Six (6) board-certified pathologists who have demonstrated proficiency in reading steatohepatitis cases, have liver subspecialty experience and sign out MASH cases in their clinical practice participated in this study. All pathologists were trained on the study protocol prior to study start.

The GT reads were collected on glass slides using a light microscope by a group of 3 pathologists. Each of these pathologists read 160 cases on glass once.

A different set of 3 pathologists performed the study reads. They read all cases twice, first utilizing WSIs on the digital pathology platform and after a 2-week washout, on glass slides with light microscopy.

### Statistics and bioinformatics

The primary endpoint was to evaluate for non-inferior overall percent agreement (OPA) of individual pathologist’s steatohepatitis evaluation (defined as NAS ≥ 4 with a score of ≥ 1 for each feature and absence of atypical features suggestive of other liver disease) on the WSI with glass GT as compared to the OPA of their glass reads with glass GT with a 0.05 non-inferiority margin. Bootstrap 95% confidence intervals and p-values were also computed. The 0.05 non-inferiority margin utilized in this study was chosen, based upon those WSI platform validations for primary diagnosis, where a non-inferiority margin of 0.04 is often used (5,6). However, inter- and intra-reader variability is higher in a complex condition like steatohepatitis and therefore non-inferiority margin of 0.05 was utilized.

The glass GT consensus score was determined as the mode if at least 2 out of 3 pathologists were in agreement. If there was no agreement, the GT was considered to be the median of all 3 scores. Additionally, the majority of the 3 GT pathologists’ responses were used to assess presence of atypical features.

The GT median scores were computed using the following method:

- For scores
  - If the median for the scores was an integer, that was the final score.
  - If the median for the scores was not an integer, analysis was performed with rounding up a score/stage, then again rounding down a score/stage, and the average of the two was used.
- For presence of atypical features (yes/no)
  - If at least 2 GT pathologists agreed, that was the final answer.
  - If at least 2 GT pathologists did not agree, analysis was performed with the answer yes, then again with the answer as no, and the average of the two was used.

Any case where 2 out of the 3 glass GT pathologists indicated either H&E or Masson’s trichrome slide was not evaluable for scoring, the whole case was removed from data analysis and if possible, was replaced with a new case, which fulfilled the target inclusion criteria. Eighteen (18) cases were deemed inadequate by the GT panel and 17 of the cases were replaced. If a study pathologist indicated that any slide (either H&E or Masson’s trichrome) was not adequate for scoring, that slide was removed from data analysis for that pathologist only.

The secondary endpoint consisted of study pathologist scores for the three steatohepatitis features, CRN fibrosis and the overall NAS score between WSI and glass read. This endpoint was evaluated as follows: Linearly WK concordance statistics between glass and WSI read for each of the pathologists (intra- pathologist, inter-modality), each of the 4 histologic features (steatosis, lobular inflammation, hepatocellular ballooning and fibrosis), and overall NAS score. Overall, linearly WK was computed for each feature and overall NAS score by averaging the WK for the 3 pathologists. Bootstrap 95% confidence intervals are provided on the overall linearly WK. These concordance estimates are compared to the published range in Table 1. These analyses are based on observed data.

**Table 1.**
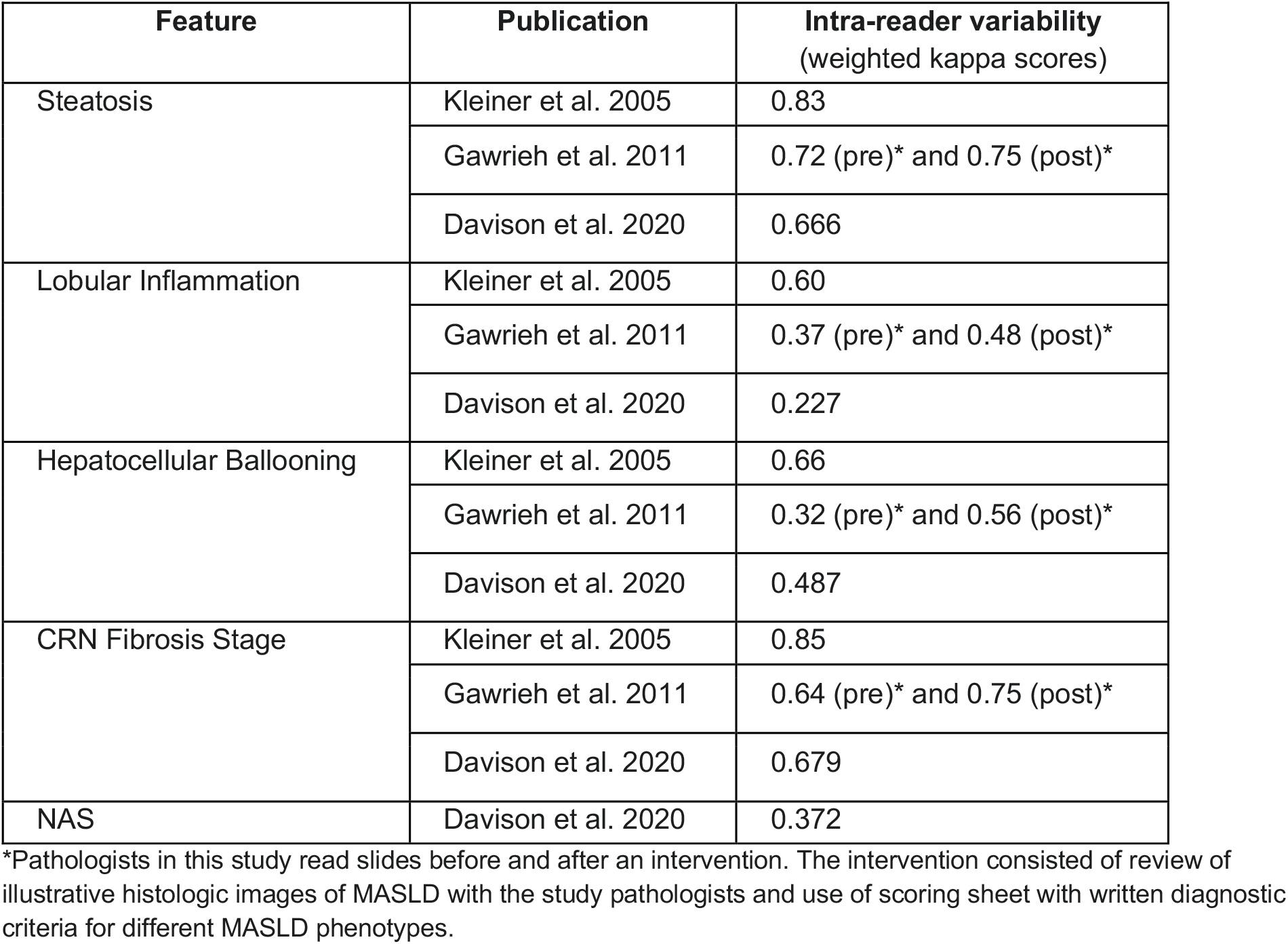
WK scores for intra-reader variability.

The exploratory endpoint determined overall WKs for steatosis, hepatocellular ballooning, lobular inflammation fibrosis and NAS for WSI with glass GT as compared to the WKs for glass reads with glass GT.

### Determination of Sample Size

The 2022 CAP updated guidance for validating WSI systems (22) for pathology applications, recommends using a sample set of at least 60 cases based on evidence from 33 publications reviewed. Based on these 33 studies, CAP also recommends the ideal validation study endpoint is 95% intra-rater diagnostic concordance between digital and glass slides. However, they note that non-inferiority design is also acceptable. CAP guidelines though are directed towards top-line primary diagnosis, and following the guidelines for disease states and scoring systems where intra-rater variability is high (e.g., steatohepatitis) can be challenging. Given the published variability, both inter- and intra-reader, 60 case datasets might not be enough to be statistically powered for any significant conclusions. Given the ranges of expected variability in expert reads for diagnosis and scoring of steatohepatitis, a sample size of 160 slides was selected to provide a degree of precision around the estimates, to account for non-evaluable slides, and any incidental breakage of glass slides.

## Results

One hundred and fifty-nine (159) cases were enrolled in the study by 3 GT pathologists by reading glass slides using a light microscope. Distribution of slides based on slide level score from glass GT are listed in Table 2. Based on the study glass GT, the slide set included 38.9% of cases that met the definition for challenging, borderline cases (defined as NAS ≥4 with a score of 0 for at least one of the features, NAS =4 with a score of ≥1 for each of the features or NAS =3).

**Table 2.**
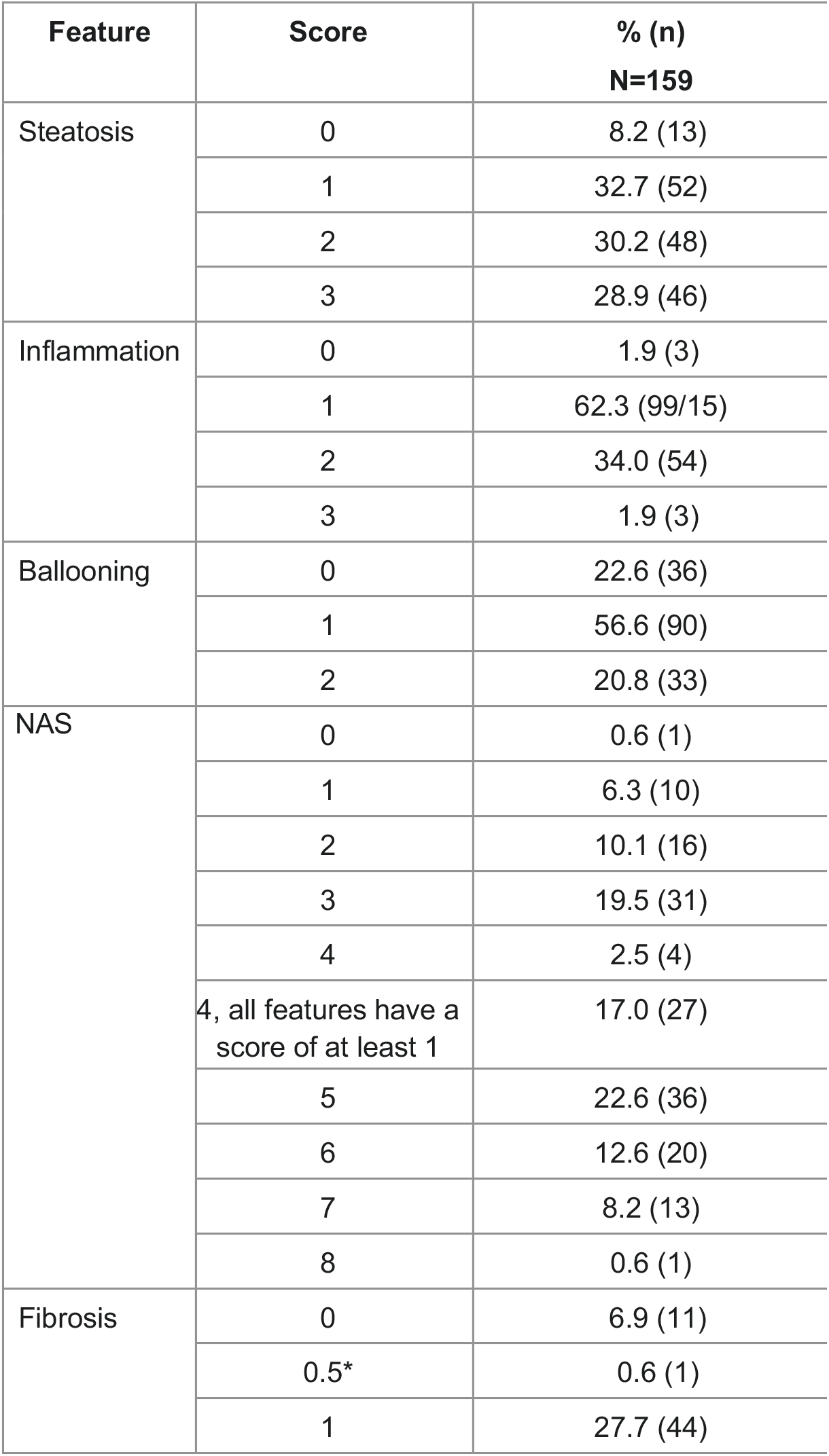
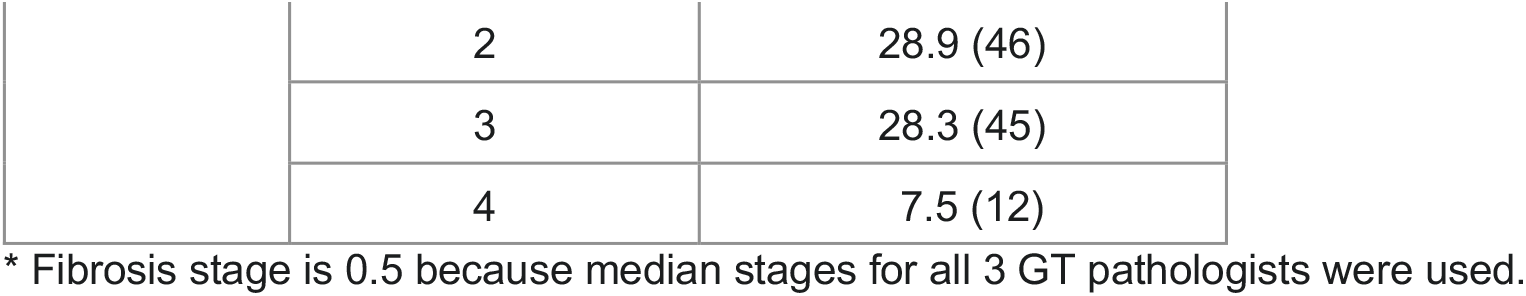
Distribution of slides based on glass GT.

Overall, the 3 study pathologists indicated presence of atypical features 57 times and for 40 of these, the categorization was identical on glass and on WSI. For the discrepant cases, the WSI agreed with the GT 10 times and glass agreed with GT 6 times (list of atypical features in Table S1).

The acceptance criteria for non-inferiority (with a margin of 0.05) agreement for steatohepatitis evaluations between reads on WSI and glass GT compared to reads on glass and glass GT was met with a difference of -0.001 (95% CI, -0.027, 0.026; p<0.0001; Table 3). The agreement between study pathologists reads on WSIs and glass GT was 0.743 (95% CI, 0.7, 0.788) and the agreement between glass reads and glass GT was 0.745 (95% CI, 0.703, 0.786).

**Table 3.** OPA between reads on WSI and glass GT vs reads on glass and glass GT.

| Modality | N | Agreement Rate (95% CI) | Difference (95% CI) | P-value |
| --- | --- | --- | --- | --- |
| WSI vs GT | 159 | 0.743 (0.7, 0.788) | -0.001 (-0.027, 0.026) | <0.0001 |
| Glass vs GT | 159 | 0.745 (0.703, 0.786) |  |  |

Agreement for steatohepatitis evaluations between reads on WSI and glass GT compared to reads on glass and glass GT were similar for all 3 pathologists (Table 4). For pathologist A, the difference between WSI reads and glass GT vs glass reads and glass GT was -0.006 (95% CI, -0.031, 0.0196). For pathologist B the difference between WSI reads and glass GT vs glass reads and glass GT was 0.0278 (95% CI, -0.034, 0.089) and the difference for pathologist C was -0.025 (95% CI, -0.069, 0.016).

**Table 4.** OPA between reads on WSI and glass GT vs reads on glass and glass GT by individual pathologist.

| Pathologist | Modality | N | Agreement Rate (95% CI) | Difference (95% CI) |
| --- | --- | --- | --- | --- |
| A | WSI vs GT | 159 | 0.843 (0.786, 0.899) | -0.006 (-0.031, 0.019) |
|  | Glass vs GT | 159 | 0.849 (0.792, 0.906) |  |
| B | WSI vs GT | 158 | 0.633 (0.56, 0.707) | 0.0278 (-0.034, 0.089) |
|  | Glass vs GT | 157 | 0.605 (0.529, 0.679) |  |
| C | WSI vs GT | 159 | 0.755 (0.686, 0.824) | -0.025 (-0.069, 0.016) |
|  | Glass vs GT | 159 | 0.780 (0.711, 0.843) |  |

WKs between WSI read and glass read for each steatohepatitis feature and CRN fibrosis (Table 5) overall and per pathologist (intra-reader, inter-modality agreements) were also determined (Table 6). For each histologic feature, the overall WKs were higher than published values (Table 1). For features per individual pathologist, pathologist A and C had the highest WKs across all features and these WKs were all higher than published intra-pathologist WKs (Table 1). For pathologist B, the WKs were lower than pathologist A and C but still in the published ranges listed in Table 1. For overall NAS score, all 3 pathologists were higher than the published values from Davison 2020 (23) (Table 1), with WKs being the highest for pathologist A and C.

**Table 5.** Average WK between WSI reads and glass reads per histologic feature.

| Feature | N | Weighted Kappa (95%CI) |
| --- | --- | --- |
| Steatosis | 159 | 0.882 (0.844, 0.916) |
| Inflammation | 159 | 0.761 (0.707, 0.809) |
| Ballooning | 159 | 0.788 (0.732, 0.835) |
| Fibrosis | 159 | 0.872 (0.837, 0.901) |
| NAS | 159 | 0.795 (0.76, 0.825) |

**Table 6.** WK between WSI reads and glass reads per histologic feature by pathologist.

| Pathologist | Feature | N | Weighted Kappa (95% CI) |
| --- | --- | --- | --- |
| A | Steatosis | 159 | 0.987 (0.966, 1) |
|  | Inflammation | 159 | 0.938 (0.888, 0.981) |
|  | Ballooning | 159 | 0.926 (0.862, 0.972) |
|  | Fibrosis | 159 | 0.941 (0.903, 0.972) |
|  | NAS | 159 | 0.956 (0.93, 0.977) |
| B | Steatosis | 157 | 0.741 (0.635, 0.83) |
|  | Inflammation | 157 | 0.545 (0.441, 0.645) |
|  | Ballooning | 157 | 0.618 (0.498, 0.719) |
|  | Fibrosis | 153 | 0.733 (0.654, 0.803) |
|  | NAS | 157 | 0.602 (0.524, 0.668) |
| C | Steatosis | 159 | 0.918 (0.87, 0.962) |
|  | Inflammation | 159 | 0.801 (0.716, 0.877) |
|  | Ballooning | 159 | 0.821 (0.746, 0.885) |
|  | Fibrosis | 158 | 0.942 (0.905, 0.972) |
|  | NAS | 159 | 0.828 (0.779, 0.874) |

WKs between WSI read and glass GT compared to glass read and glass GT for each steatohepatitis feature and CRN fibrosis (Table 7) were determined. For each histologic feature, the overall WKs were comparable between WSI read vs GT and glass vs GT with overlapping confidence intervals for all histologic features.

**Table 7.** WK between reads on WSI and glass GT vs reads on glass and glass GT for each histologic.

| Feature | Modality | N | Weighted Kappa (95% CI) |
| --- | --- | --- | --- |
| Steatosis | WSI vs GT | 159 | 0.58 (0.505, 0.64) |
|  | Glass vs GT | 159 | 0.593 (0.519, 0.655) |
| Inflammation | WSI vs GT | 159 | 0.367 (0.3, 0.432) |
|  | Glass vs GT | 159 | 0.38 (0.315, 0.445) |
| Ballooning | WSI vs GT | 159 | 0.537 (0.457, 0.608) |
|  | Glass vs GT | 159 | 0.522 (0.435, 0.595) |
| Fibrosis | WSI vs GT | 159 | 0.64 (0.574, 0.695) |
|  | Glass vs GT | 159 | 0.604 (0.536, 0.662) |
| NAS | WSI vs GT | 159 | 0.527 (0.476, 0.573) |
|  | Glass vs GT | 159 | 0.525 (0.473, 0.571) |

## Discussion

This digital pathology platform validation study demonstrates that the accuracy of steatohepatitis digital reads on the WSI viewer is equivalent to reads performed with traditional light microscopy with glass slides. Additionally, the agreement between WSI and glass GT reads vs glass and glass GT reads were shown to be similar for each individual participating pathologist. Use of a digital pathology platform facilitates multiple independent pathologists reads in parallel and in consensus sessions, as is now commonly performed for MASH trials and recommended by the FDA. This digital and glass read equivalence is in line with studies performed for primary diagnoses by Leica (6) and Philips (5). This study demonstrated a significant non-inferior OPA of steatohepatitis assessment between WSI and glass GT reads vs glass and glass GT reads (NI margin of 0.05, difference of -0.001, 95% CI of (−0.027,0.026), and p<0.0001). Additionally, the OPA between WSI and glass GT reads vs glass and glass GT reads were shown to be similar for each individual participating pathologist. Average intra-reader, inter-modality WKs for each histologic score feature in this study were higher than WKs in published literature (Table 1).

Varying level of intra-reader agreement was observed per pathologist per histologic feature, which is expected, as a wide range of intra-reader WKs have been demonstrated in the literature (23–25). Importantly, results from all 3 pathologists were within the published ranges for intra-reader WKs, with 2 out of the 3 pathologists exceeding the published WKs for all 4 histologic features (steatosis, lobular inflammation, hepatocellular ballooning and fibrosis) and all 3 pathologists exceeding the WK for overall NAS score.

To ensure that the platform was validated across steatohepatitis spectrum, the target study population was intended to be enriched with 5-10% challenging or borderline steatohepatitis cases (defined as NAS ≥ 4 with a score of 0 for at least one of the features, NAS = 4 with a score of ≥1 for each of the features or NAS = 3). Previously collected single central pathologist scores were used during study enrollment; however, based on the final study consensus GT, around 40% of the cases met the definition of being borderline or challenging. The observed difference in target vs. actual percent challenging enrichment is consistent with published literature describing inter-pathologist agreement rates for NAS of approximately only 30% (23). This level of enrichment is in contrast to primary diagnosis studies where only around 5% of the cases included were considered to be borderline and/or challenging, and only major discordances in categorical diagnoses were counted towards disagreement rates, whereas any histologic feature score discrepancy was considered here. However, even with ~40% challenging cases in this study, the 0.05 non-inferiority margin was met.

The results from this validation study support the conclusion that the digital pathology platform is equivalent to the glass read in reference to a robust glass GT, when used by pathologists to evaluate steatohepatitis trial populations for diagnosis and trial-based scoring criteria (defined as NAS ≥ 4 with a score of ≥ 1 for each feature and absence of atypical features suggestive of other liver disease) and histologic feature scoring during enrollment and for follow-up timepoints. This digital pathology platform, along with the Leica AT2 WSI scanner, can therefore be utilized for individual and consensus steatohepatitis reads in clinical trials. The work presented here was designed specifically for use in clinical trials, utilizing a robust design with MASH trial-specific endpoints in a challenging study population, representative of both screened and enrolled patient samples. Incorporating digital pathology into clinical trial workflows makes trial management more efficient, allows for multiple reads in parallel, and provides opportunities to utilize the most experienced pathologists on reader panels as geographic location is no longer a factor for selecting pathologists or shipping glass slides. Utilization of a digital pathology platform will allow pathologists from all over the world to work on the same cases simultaneously and provide their results within hours of slide upload, shortening trial timelines, while allowing for accurate, gold standard assessments.

## Data Availability

All data produced in the present study are available upon reasonable request to the authors

### Appendix

**Table S1.**
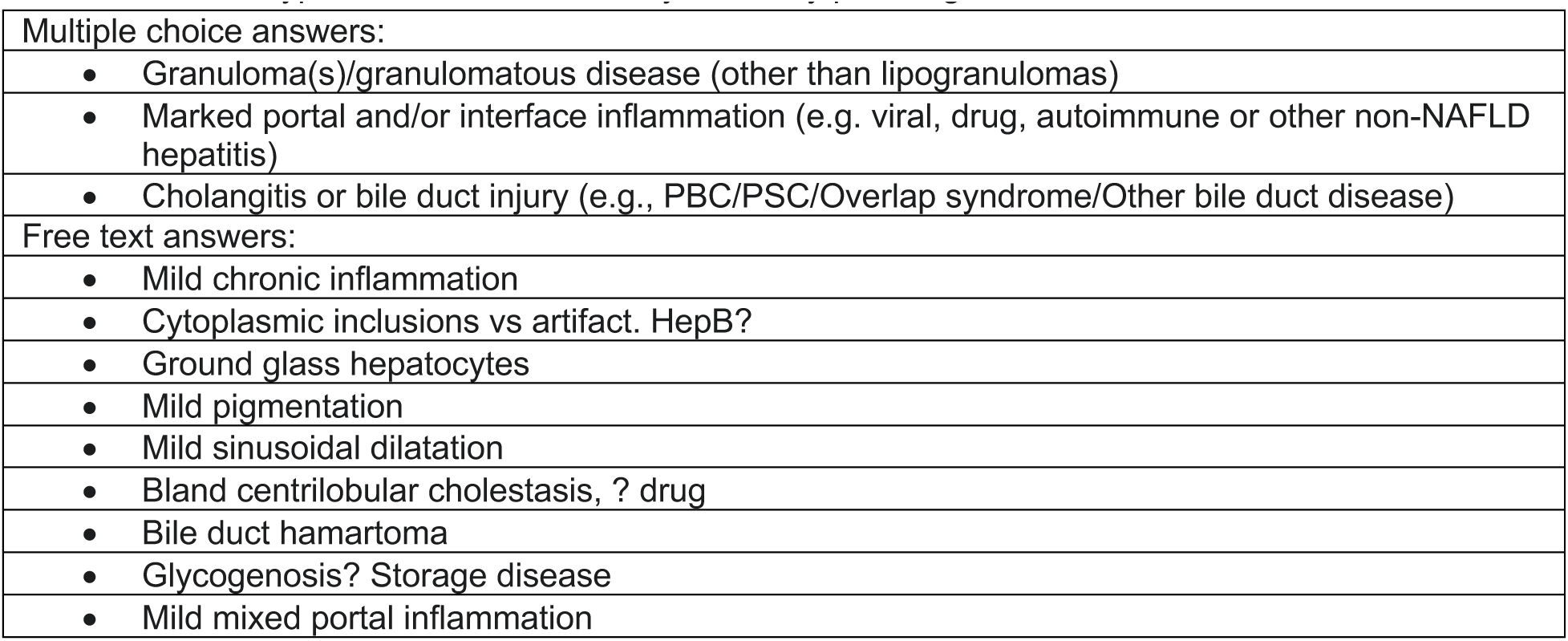
List of atypical features as noted by the study pathologists.

